# Chondrocalcinosis and the haemochromatosis linked *HFE* C282Y homozygous variant in the UK Biobank

**DOI:** 10.1101/2025.03.18.25324165

**Authors:** Lucy R. Banfield, Karen M. Knapp, Luke C. Pilling, David Melzer, Janice L. Atkins

**Author notes:** <u>Corresponding author:</u> Dr Janice L Atkins, College House, St. Luke’s Campus, University of Exeter, Exeter, Devon, EX1 2LU, UK.

## Abstract

**Background:** C282Y genetic homozygosity is the main cause of the iron-overload disorder haemochromatosis. Musculoskeletal pain and arthropathy are common in haemochromatosis, but less is known about chondrocalcinosis (cartilage calcification) with the C282Y variant, especially in the community. We assessed knee chondrocalcinosis in iDXA (dual-energy X-ray absorptiometer) images from UK Biobank volunteers by *HFE* genotype.

**Methods:** Data were from 236 European genetic ancestry C282Y homozygotes and 236 age, sex, and BMI-matched controls with no C282Y alleles (48-80 years, mean 64.6, SD ±7.6). 435 of 472 participants had relevant left and right knee iDXA imaging. Evidence of chondrocalcinosis was assessed by an experienced reporting radiographer blind to genotype to ensure unbiased and objective evaluation. Logistic regression models were age, sex and BMI matched.

**Results:** Knee chondrocalcinosis was present in 15.9% (14/88) of C282Y homozygous males and <5.9% (<5/84) of males without variants (OR=7.76, 95% CI: 1.71–35.25, p=0.008). 57.1% (8/14) of the male homozygotes with knee chondrocalcinosis reported knee pain during the previous three months, but <28.6% also had a haemochromatosis diagnosis. In females, 6.0% (8/134) of C282Y homozygotes had knee chondrocalcinosis, vs <3.9% (<5/129) without variants. However, the odds of chondrocalcinosis were not significantly higher in C282Y homozygotes (OR=1.98, 95% CI: 0.58-6.76, p=0.273), therefore a larger sample size may be required to detect a smaller effect in female homozygotes.

**Conclusion:** In this community genotyped sample, male C282Y homozygotes had a markedly increased odds of knee chondrocalcinosis. Evaluation of serum ferritin levels to identify possible haemochromatosis may be justified in knee chondrocalcinosis management.

## Introduction

Haemochromatosis is an autosomal recessive condition resulting in iron overload predominantly caused by mutations in the *HFE* gene, in particular the C282Y mutation and to a lesser extent the H63D (1). C282Y homozygosity is the mutation primarily associated with haemochromatosis, compared to other lower penetrance C282Y/H63D genotypes (1). Consequently, for the purpose of this study we elected to focus on C282Y homozygotes.

This excessive absorption of iron associated with haemochromatosis and its subsequent accumulation, can have a detrimental impact on several different body organs. The clinical disease has been associated with osteoarthritis (OA), diabetes, liver disease including liver cancer, as well as several other conditions(2–4) Despite being the most common genetic disease in those of Northern European ancestry, diagnoses of haemochromatosis may be missed or delayed, with only a minority of community populations with the C282Y homozygote variant being diagnosed(5).

There are several musculoskeletal impacts that have been associated with haemochromatosis in those with the C282Y homozygous mutation and particularly among males. A number of previous studies have identified increased risk of joint replacement surgeries(5–8), osteoporosis and fracture(9–12). The arthritis of haemochromatosis has been a well-recognised component of the disease for several years with findings typically being described as affecting primarily the metacarpophalangeal joints (MCPJs), the ankle, knee and hip, but also other less typical joints such as shoulders and wrists (13– 15). Clinical presentation can include pain, swelling, stiffness and a limited range of movement with exuberant osteophyte formation. These findings can often be bilateral and are reported to occur in two-thirds of those with haemochromatosis(16,17).

The mechanism by which iron overload causes this degenerative process is still not entirely clear, but one hypothesis is that the iron overload and excessive haemosiderin deposition in the synovial membrane may cause inflammation of the synovial tissues, articular cartilage and other structures within the joint resulting in degeneration. A study by Dejaco et al identified that inflammatory changes and synovitis were prevalent in those with haemochromatosis arthropathy, and that subclinical inflammation was also seen in the absence of haemochromatosis arthritis(18). This is supported by a recent study which identified that iron overload may induce M1 polarisation in macrophages resulting in the release of pro-inflammatory factors and resulting in joint degeneration(19) Macrophages are one of the two main cellular components of synovium and serve to engulf pathogens and debris relative to ‘normal’ ageing of tissues to maintain a healthy joint(20). However, if abnormally activated, M1 macrophage pro-inflammatory cytokines are the key drivers of chronic inflammation and can also contribute to tissue damage, and while this can be beneficial in the short term when fighting infections, it can be detrimental if chronic.

With regards to chondrocalcinosis (CC, cartilage calcification), there is wide variation in the reported frequency within haemochromatosis patients, with estimates ranging from 5 to 49% (21–23). In a small study of 18 diagnosed haemochromatosis patients undergoing treatment via venesection, CC prevalence increased from 39% to 72% during a 10-year follow-up(24) However, less is known about the incidence of chondrocalcinosis in those with the C282Y mutation. This study examines associations between the presence of chondrocalcinosis in the knee and *HFE* C282Y homozygosity (compared to age, sex, and BMI matched controls without C282Y alleles), within a subset of UK Biobank community dwelling participants.

## Methods

### UK Biobank participants

UK Biobank (UKB) is a cohort study of ∼502,150 UK adults aged 37-73 years at baseline assessment (2006 to 2010). Data includes baseline characteristics, biomarkers, genetics, and linked medical records. UKB data are available to any bone fide researcher following application. The Northwest Multi-Centre Research Ethics Committee approved the collection and use of UKB data (Research Ethics Committee reference 11/NW/0382). Participants gave informed consent for the use of their data, health records, and biological materials for health-related research purposes. Access to UKB was granted under application number 14631.

### Genotype data

Genotype information on *HFE* C282Y (rs1800562 A allele) and *HFE* H63D (rs1799945 G allele) were from whole exome sequencing data (methods by Regeneron(25)). Patient consent did not include feedback to patients on genotypes. We included 451,270 participants genetically similar to the 1000 genome project European Ancestry superpopulation (‘EUR-like’(26)), of which 2,902 (0.64%) were C282Y homozygotes.

### iDXA (dual-energy X-ray absorptiometer) data

During follow-up, a subset of UK Biobank participants (n=41,594/451,270 at the time of analysis) attended a follow-up visit for iDXA imaging (2014 to 2020, aged 45 to 82 years). As part of a suite of imaging examinations, iDXA high resolution imaging of each knee was performed using a GE Lunar (Madison, WI) (27). Within this iDXA subset, we used all available data from 236 C282Y homozygotes, along with 236 age, sex, and BMI-matched controls with no C282Y or H63D genotypes (n=472). Within these 472 participants, 435 had relevant right and left knee iDXA imaging (263 females and 172 males) (see Supplementary Table 1 for flowchart of included participants). A small number of this subset had undergone knee replacement surgery prior to imaging which excluded those joints from iDXA. Others were excluded due to poor image quality or missing images. These iDXA knee images were reviewed for radiological evidence of chondrocalcinosis within the tibiofemoral joint space by an experienced reporting radiographer (LRB), blind to participant genotype to ensure unbiased and objective evaluation. Chondrocalcinosis (CC) was classified if present in either the left or right knee (see Figure 1).

**Figure 1.**
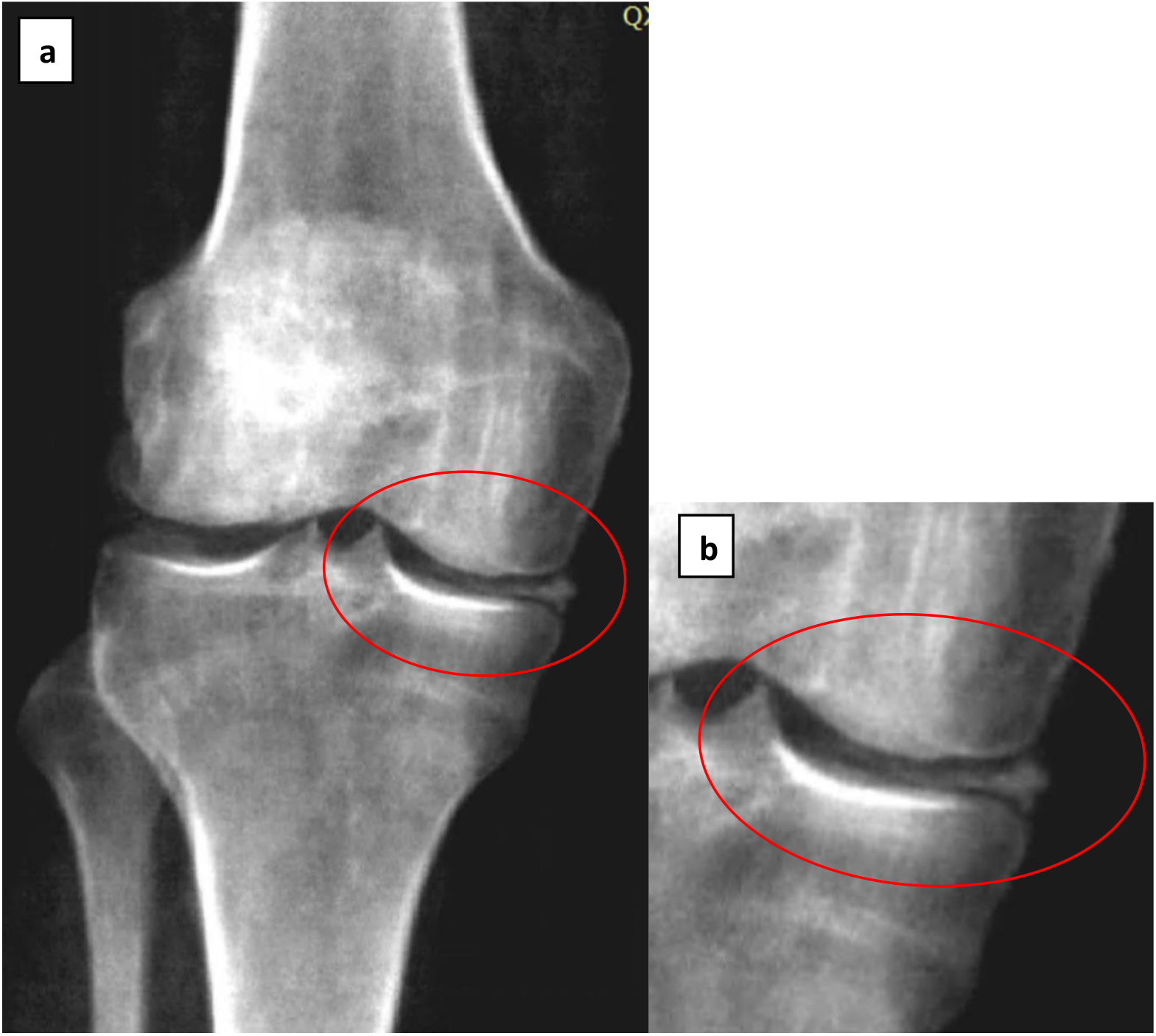
Example of chondrocalcinosis of the knee on an iDXA image in UK Biobank (a) with an enlarged picture (b) to highlight the presence of chondrocalcinosis in the medial tibiofemoral space. Reproduced by kind permission of UK Biobank©

We excluded participants who had ever had a diagnosis of rheumatoid arthritis, multiple myeloma, malignant neoplasm and secondary malignant neoplasm of bone, also benign neoplasm of bone (including diagnoses from baseline self-report and hospital inpatient follow-up until 2022) (see Supplementary Table 2 for coding used).

### Additional variables (at the time of the iDXA)

Participants were asked to self-report if they had knee pain that lasted more than 3 months. The prevalence of haemochromatosis and OA was based on diagnoses from baseline self-reports plus hospital inpatient data from 1996 to the date of the iDXA visit, as coded previously (7).

### Statistical analysis

All analyses were carried out in STATA 18.0 using a matched analysis. *HFE* C282Y homozygotes were matched to controls (without C282Y or H63D alleles) for sex, age and body mass index (BMI) category at time of DXA - underweight (<18.5kg/m^2^), normal (18.5-24.9kg/m^2^), overweight (25-29.9kg/m^2^) or obese (≥30kg/m^2^). We used random-effects logistic regression models to estimate associations between genotype and likelihood of knee chondrocalcinosis using STATA function `xtlogit`, specifying pairwise matching as the random intercept to account for the within-pair correlation. This approach was chosen due to the matched case-control design of our study. The xtlogit model is particularly suitable for or analysing matched pair data with binary outcomes, effectively accounting for within-subject correlation and unobserved heterogeneity through random-effects models.(28)

We examined the prevalence of chronic knee pain and OA within participants by genotype group and the presence of chondrocalcinosis. We also performed additional sensitivity analysis, excluding those diagnosed with haemochromatosis at the time of the iDXA imaging.

## Results

### Characteristics of participants

Analyses included 435 European descent participants aged 48 to 80 years at the time of iDXA scanning, 60.5% of the sample were women (n=263) and the mean age of the cohort was 64.6 years (SD ±7.6). There were 222 C282Y homozygotes (134 females and 88 males) (Table 1).

**Table 1.** UK Biobank participant characteristics of C282Y homozygotes with available iDXA data and randomly selected matched controls, with no *HFE* C282Y or H63D alleles*.

| Group |  | *HFE*<br>genotype | Mean age,<br>years (SD) | Haemochromatosis<br>diagnosis at time<br>of DXA (%) | OA at<br>time of<br>DXA (%) | Knee pain<br>for 3+<br>months (%) |
| --- | --- | --- | --- | --- | --- | --- |
| **Males** | **No CC** | No C282Y or H63D<br>alleles | 64.2 (8.0) | 0 (0) | 8 (9.8) | 8 (9.8) |
|  |  | C282Y homozygotes | 64.1 (8.0) | 20 (27.0) | <5 (<6.8) | 13 (17.6) |
|  | **CC**<br>**present** | No C282Y or H63D<br>alleles | 72.5 (3.5) | 0 (0) | 0 (0) | <5 (<50.1) |
|  |  | C282Y homozygotes | 66.6 (7.3) | <5 (<35.7) | 0 (0) | 8 (57.1) |
| **Females** | **No CC** | No C282Y or H63D<br>alleles | 64.4 (7.3) | 0 (0) | 14 (11.2) | 15 (12.0) |
|  |  | C282Y homozygotes | 64 .6 (7.6) | 12 (9.5) | 10 (7.9) | 23 (18.3) |
|  | **CC**<br>**present** | No C282Y or H63D<br>alleles | 73.5 (4.5) | 0 (0) | 0 (0) | 0 (0) |
|  |  | C282Y homozygotes | 68.0 (7.4) | 0 (0) | 0 (0) | <5 (<62.5) |

\*Controls matched on age, sex and BMI category.
Abbreviations: CC, chondrocalcinosis; OA osteoarthritis.

Overall, CC was present in either (or both) knees of 28 (6.4%) participants, 22 of which were C282Y homozygotes. In males, knee CC was present in 15.9% of C282Y homozygotes (14/88) and <5.9% without *HFE* variants (<5/84). In females, knee CC was present in 6.0% of C282Y homozygotes (8/134) and <3.9% without *HFE* variants (<5/129). The mean age of those with CC was lower for both the male and female C282Y homozygotes (66.6 years and 68.0 years respectively) when compared to matched controls with no *HFE* mutations (72.5 years and 73.5 years).

Matched regression analysis demonstrated significantly increased odds of CC in the C282Y homozygotes (Odds Ratio: 3.79, 95% Confidence Intervals: 1.51 – 9.55, p=0.005), compared to age, sex and BMI-matched controls without *HFE* C282Y or H63D alleles. The same analysis stratifying by sex, demonstrated increased odds for male homozygotes (OR: 7.76, 95% CI: 1.71 – 35.25, p=0.008), and although a modest increase in odds was also seen in female homozygotes, this was not significant (OR: 1.98, 95% CI: 0.58-6.76, p=0.273) (Table 2).

**Table 2.** Increased odds of chondrocalcinosis in HFE C282Y homozygotes compared to controls*.

| Group | HFE genotype | N | N CC (%) | OR | 95% CIs | p-value |
| --- | --- | --- | --- | --- | --- | --- |
| **Males** | No C282Y or H63D alleles | 84 | <5 (<5.9) | 1.00 |  |  |
|  | C282Y homozygotes | 88 | 14 (15.9) | 7.76 | 1.71–35.25 | 0.008 |
| **Females** | No C282Y or H63D alleles | 129 | <5 (<3.9) | 1.00 |  |  |
|  | C282Y homozygotes | 134 | 8 (6.0) | 1.98 | 0.58–6.76 | 0.273 |
| **All** | No C282Y or H63D alleles | 213 | 6 (2.8) | 1.00 |  |  |
|  | C282Y homozygotes | 222 | 22 (9.9) | 3.79 | 1.51-9.55 | 0.005 |
|  | *Total* | 435 | 28 (6.4) |  |  |  |

\*Controls with no HFE C282Y or H63D alleles, matched on age, sex and BMI category.
Abbreviations: CC, chondrocalcinosis. OR, Odds Ratio. CIs, Confidence Intervals.

### Sensitivity analyses (excluding haemochromatosis diagnosis)

To determine whether clinical haemochromatosis impacted on CC prevalence within this subset we excluded those who had been diagnosed with haemochromatosis at the time of iDXA imaging. Within our dataset there were 36 homozygotes with a haemochromatosis diagnosis (n=24 male, 27.3%, and n=12 female, 9.0%). When those with a haemochromatosis diagnosis at time of imaging (n=36) were excluded, a higher proportion of CC was still evident in both male (n=10, 15.6%) and female (n=8, 6.6%) homozygotes when compared to the wildtype male (<5.9%) and female (<3.9%) participants. This increased risk remained high for male homozygotes who had not been diagnosed with haemochromatosis at the time imaging (OR=7.59, 95% CI: 1.60-35.99, p=0.011), but again, although increased odds were seen in the female homozygotes, this was not significant (OR=2.19, 95% CI: 0.64-7.48, p=0.210) (Table 3).

**Table 3.** Increased odds of chondrocalcinosis in HFE C282Y homozygotes compared to controls, excluding those with diagnosed haemochromatosis*.

| Group | HFE genotype | N | N CC (%) | OR | 95% CIs | p-value |
| --- | --- | --- | --- | --- | --- | --- |
| **Males** | No C282Y or H63D alleles | 84 | <5 (<5.9) | 1.00 |  |  |
|  | C282Y homozygotes | 64 | 10 (15.6) | 7.59 | 1.60-35.99 | 0.011 |
| **Females** | No C282Y or H63D alleles | 129 | <5 (<3.9) | 1.00 |  |  |
|  | C282Y homozygotes | 122 | 8 (6.6) | 2.19 | 0.64-7.48 | 0.210 |

\*Controls with no HFE C282Y or H63D alleles matched on age, sex and BMI category.
Abbreviations: CC, chondrocalcinosis.

### Osteoarthritis and joint replacement associations

The prevalence of OA at the time of DXA imaging was slightly higher in those without *HFE* mutations (12 homozygotes vs. 22 wildtypes), and of these, a greater number was observed in female (n=10) homozygotes compared to male (n=<5) homozygotes. No homozygous participants had concurrent OA and CC at the time of imaging.

### Knee pain

When investigating the association between “knee pain for 3+ months” and the presence of CC, we again found reported pain and CC to be more prevalent among male homozygotes (n=8, 57.1%) compared to those without the *HFE* mutation (<50%). Conversely, the number of female participants with both CC and reported “knee pain for 3+ months” was notably lower, with only one female homozygote and no wildtype females affected (Table 1). Excluding individuals diagnosed with haemochromatosis reduced the number of male homozygotes with both pain and CC further.

## Discussion

We used a subset of participants from the UK Biobank cohort study (the largest community sample of haemochromatosis-genotype individuals to date) to examine associations between *HFE* C282Y homozygosity and the presence of chondrocalcinosis within the tibiofemoral joint as identified on iDXA imaging. We specifically focused on the high risk C282Y homozygotes for this study, excluding other *HFE* related mutations such as H63D homozygotes and C282Y-H63D compound heterozygotes. We compared the C282Y homozygotes to age, sex, and BMI-matched controls without *HFE* genotypes. Our analysis revealed a higher incidence of chondrocalcinosis in the knee joints of both male and female C282Y homozygotes compared to matched wildtypes, with a notably elevated risk observed among male homozygotes. While previous studies have reported increased prevalence of chondrocalcinosis associated with clinical haemochromatosis, our findings offer a more detailed understanding of the genotype-specific risk within a large community-based sample of predominantly undiagnosed individuals, and to our knowledge is the largest study to date on community genotyped C282Y homozygotes and CC.

CC within the knee joint is commonly associated with degenerative changes and is frequently seen in conjunction with osteoarthritis (OA) (29,30) but can also be seen in several other conditions including hyperparathyroidism, hypophosphatasia as well as haemochromatosis. (31) A previous paper by Timms and colleagues recruited participants according to the presence of either CC or calcium pyrophosphate dihydrate disease and then subsequently assessed for associations via genotype and with control participants (n=18 C282Y homozygotes). Although this analysis resulted in a small sample of C282Y homozygotes, and their results were not stratified by sex in comparison to our study, they did identify a connection between C282Y homozygosity and chondrocalcinosis (relative risk 3.4, p=0.037).(32) Our findings further confirm the significant associations between homozygosity and tibiofemoral chondrocalcinosis when compared to those with no *HFE* genetic mutations and in addition highlights the incidence of CC in male homozygotes is almost double that of the female homozygotes.

Our data analysis also identified that the mean age of those within our study who were C282Y homozygotes and had tibiofemoral CC, was lower than the wildtypes. This is in common with other studies looking at joint changes seen with haemochromatosis. (33,34) The participants in these studies have usually been those with a haemochromatosis diagnosis, however when we excluded those in our cohort who had been diagnosed at the time of iDXA, we continued to see a lower mean age within the C282Y homozygotes.

These findings suggest that chondrocalcinosis in this context is relative to the *HFE* mutation and metabolic in nature, although the extent of iron loading in these participants is unknown. Whereas the CC observed in those with no *HFE* mutations is more likely to be attributable to age related changes and degenerative disease. This supposition may be further strengthened by the apparent lack of OA in our cohort of homozygotes with CC, although it should be noted that the data was derived from hospital records and did not include primary care data, so numbers with OA at the time of imaging may be under reported in this study.

Chronic pain is another characteristic associated with CC, haemochromatosis and C282Y homozygosity (35,36) and although we saw increased numbers of male and female homozygotes reporting knee pain, when we assessed these outcomes against the presence of CC, we found no significant increased associations between pain, homozygosity and the presence of CC.

### Strengths and limitations

This study has several strengths. It is the largest cohort of community genotyped C282Y homozygotes, with iDXA data for the assessment of chondrocalcinosis to date. It has provided robust data in the form of imaging data, and self-report, and hospital records to evaluate several factors that could contribute to or initiate the formation of chondrocalcinosis within the knee joint. The images were reviewed by an experienced reporting radiographer who has blind to genotype. However, there are some limitations to consider. Firstly, UK Biobank participants were healthier than the general population at baseline(36), and there is a small chance of misclassification bias for self-reported disease outcomes at baseline, but this is minimised by the fact a trained nurse conducted the interviews with participants. Secondly, no data were available on ferritin concentrations or transferrin saturation, so we are unable to examine iron loading within genotype groups. Third, disease diagnosis may be an underestimate as primary care data was not used in the analysis as those records were only available for a subset of this cohort. Lastly, we studied a European ancestry population so results may not be generalizable to more diverse populations.

## Conclusion

In conclusion, these findings suggest that further investigation for iron overload, including serum ferritin and transferrin saturation levels, should be considered when chondrocalcinosis is detected on imaging, especially in younger patients and those without concurrent degenerative joint disease. Such an approach may facilitate earlier identification of iron overload, allowing for timely implementation of appropriate haemochromatosis treatment regimens.

## Data Availability

All data produced in the present work are contained in the manuscript

## Acknowledgements

This research has been conducted using the UK Biobank Resource, under application 14631. This work uses data provided by patients and collected by the NHS as part of their care and support, Copyright © (2023), NHS England. Re-used with the permission of the NHS England [and/or UK Biobank]. All rights reserved. The authors wish to thank the UK Biobank participants and coordinators for this unique dataset. We would also like to thank Professor Patrick Kiely (St Georges University Hospital NHS Trust) for his valuable and constructive suggestions during the writing of this paper. His willingness to give his time has been very much appreciated. For the purpose of open access, the author has applied [a Creative Commons Attribution (CC BY) licence] to any Author Accepted Manuscript version arising.

---

**Supplementary Figure 1.**
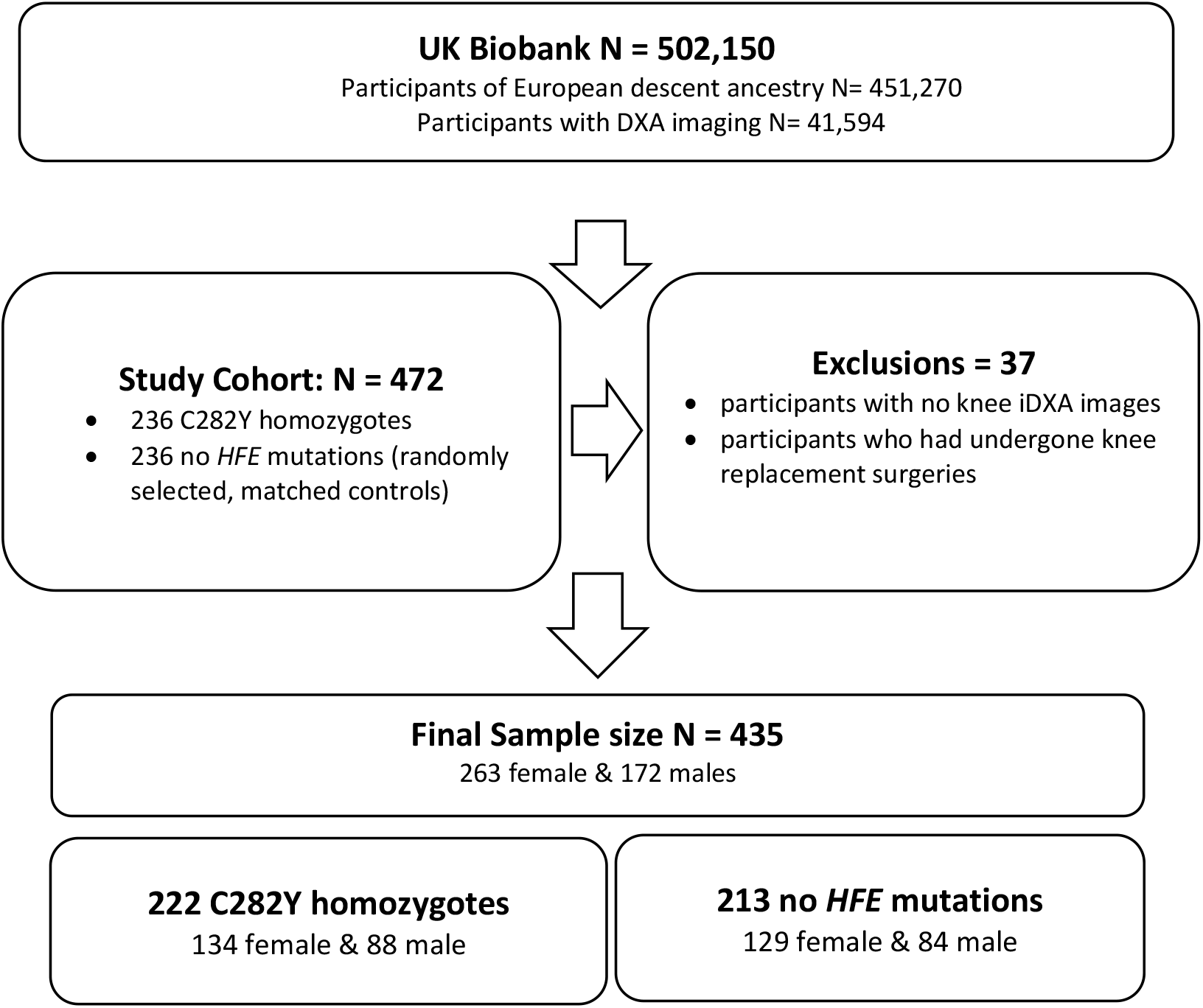
Flowchart of included/excluded participants

**Supplementary Table 1.** Incident disease coding from follow-up in hospital inpatient data

| **Disease/procedure** | **ICD-10 Codes** |
| --- | --- |
| Rheumatoid Arthritis | M05, M05.8; M05.9; M06.0; M06.8 |
| Multiple myeloma | C90.0; C90.0; C90.2; C41.0 C90.3 |
| Malignant neoplasm of bone | C40; C40.1; C40.2; C40.3; C40.8; C40.9; C41.0; C41.1; C41.2; C41.3; C41.4; C41.8; C41. |
| Secondary malignant neoplasm of bone | C79.5 |
| Benign neoplasm of bone | D16.0; D16.1; D16.2; D16.4; D16.5; D16.6; D16.7; D16.8; D16.9 |

ICD-10 = International Classification of Diseases 10<sup>th</sup> revision codes

